# Long- and short-term effects of cross-immunity in epidemic dynamics

**DOI:** 10.1101/2022.04.04.22273361

**Authors:** Iker Atienza-Diez, Luís F Seoane

**Affiliations:** Departamento de Biología de Sistemas, Centro Nacional de Biotecnología (CSIC), C/ Darwin 3, 28049 Madrid, Spain; Grupo Interdisciplinar de Sistemas Complejos (GISC), Madrid, Spain

**Keywords:** SIR, epidemiology, herd immunity, cross-immunity, covid-19, SARS-CoV-2, vaccines

## Abstract

The vertebrate immune system is capable of strong, focused adaptive responses that depend on T-cell specificity in recognizing antigenic sequences of a pathogen. Recognition tolerance and antigenic convergence cause cross-immune reactions that extend prompt, specific responses to rather similar pathogens. This suggests that reaching herd-immunity might be facilitated during successive epidemic outbreaks (e.g., SARS-CoV-2 waves with different variants). Qualitative studies play down this possibility because cross-immune protection is seldom sterilizing. We use minimal quantitative models to study how cross-immunity affects epidemic dynamics over short and long timescales. In the short scale, we investigate models of sterilizing and attenuating immunity, finding equivalences between both mechanisms—thus suggesting a key role of attenuating protection in achieving herd immunity. Our models render maps in epidemic-parameter space that discern threatening variants depending on acquired cross-immunity levels. We illustrate this application with SARS-CoV-2 data, including protection due to vaccination rates across countries. In the long-time scale, we model sterilizing cross-immunity between rolling pathogens to characterize statistical properties of successful strains. We find that sustained cross-immune protection alters the regions of epidemic-parameter space where large outbreaks happen. Our results suggest an optimistic revision concerning prospects for herd protection based on cross-immunity, including for the SARS-CoV-2 pandemics.

## I. INTRODUCTION

Responding to threats, the immune system triggers defenses of different specificity, over different time scales [1]. An *innate immunity* prompts a quick, general response against any invading pathogen. It performs non-specific tasks such as recruiting immune cells to the infection site, identifying and removing foreign substances, and initiating *adaptive immunity* by presenting antigens to T-cells. This adaptive immunity, in turn, is much more specific and long-lasting. T-cells learn to identify the presented pathogen, which will henceforth be neutralized when encountered. Memory gained by T-cells can last a life-time and is rather specific.

As a pathogen mutates, its antigenic sequences change. If this happens too quickly, strains generated within an infected host can shortly become unrecognizable by T-cells. During acute HIV infection, systemic failure occurs when too many variants overwhelm the immune system [2, 3]. A similar drifting mechanism accounts for new, yearly strains of flu virus—but its slower evolutionary pace results in new variants over the extended host population, not within each infected individual. Some cross-immunity remains depending on how much each strain has diverged from its ancestor. Historic trends of deaths due to influenza show this mechanism at work [4]: This virus evolves within a cluster of fairly similar strains for years. During such periods, the immunity gained against the cluster founder offers partial protection—decreased mortality is observed. Sporadically, the dominant strain differs enough and cross-immunity dwindles. This new, distant variant causes much larger mortality, then starts a new cluster that dominates for the next cycles.

Cross-immunity impacts epidemic dynamics at many levels. Can we describe mathematically how it alters the unfolding of two consecutive, related outbreaks? Can these effects help a population achieve herd immunity faster? Cross-reactive protection can also happen beyond immediately related strains—even across different viruses, if they share or converge upon domains of antigenic sequences [5–8]. At larger scales, how does cross-immunity change overall statistical properties of recurring epidemics? How much drift can we expect between strains that cause large pandemics? And between variants causing smaller outbreaks? How are these properties affected by a population’s lasting immune memory?

We tackle these questions using compartmental epidemiological models. The first, simplest such model, SIR, was introduced by Kermack and McKendrick in 1927 [9]. They divided a host population into: *S*, for susceptible; *I*, for infected; and *R*, for removed. Infected individuals recover spontaneously (*I →R*) at a rate *γ*. Susceptible hosts become infected (*S →I*) at a rate *β*, the pathogen’s infectivity. This model initially portrays either a decaying or rising number of infections, depending on whether the susceptible population is below or above a threshold. In the second case, as the susceptible compartment is consumed, a spontaneous decay ensues after the model’s threshold is reached—resulting in the characteristic SIR peak. In this model, individuals that moved into the recovered compartment become immunized. This is the origin of *herd immunity*—protected individuals are so abundant that the pathogen cannot find enough susceptible hosts.

This simple model illuminates the basics of epidemic dynamics, but it cannot capture the full range of phenomena showcased by infectious outbreaks (e.g. cyclic behaviors, the relevance of social structure, etc.). The SARS-CoV-2 pandemic also exposed the limitations of compartmental models to make helpful predictions [10]. Numerous alternatives are being developed to mitigate these shortages [11–18]. Several models have incorporated cross-immunity as well [5, 19–21], often mixed with additional ingredients (e.g. age structure, reinfection, seasonal forcing, etc.). These models are often used to study the dynamical coexistence of viral strains. Here, we attempt to tackle simpler questions in the most minimal models possible, with the hope of understanding cross-immunity on its own. With this, we sketch limit cases that must be recovered by more elaborate models as additional ingredients are tuned down.

In section III.A we study two consecutive SIR-like outbreaks. The first pathogen elicits a cross-immune response during the second outbreak. We study two distinct mechanisms, *sterilizing* and *attenuating cross-immunity*, finding a mathematical correspondence between them. Either mechanism alters the SIR threshold for herd immunity. We plot resulting thresholds as a function of SIR parameters and cross-immunity levels— thus visualizing conditions that prevent the second outbreak. It does not matter whether immunity is reached artificially, hence we also chart protection offered at different vaccination rates and as a function of vaccine efficacy. We showcase this useful analysis with empirical measurements form SARS-CoV-2 variants. In section III.B we study statistical properties of epidemics with sterilizing cross-immunity over longer time periods. We consider multiple SIR outbreaks, each caused by a new strain parsimoniously evolved from previous ones. To simulate this descent, we perform random walks over SIR parameter space and over the range of cross-immunity. We find that cross-immunity alters the combination of SIR parameters resulting in the largest outbreaks. We study how our results depend on population renewal, which measures immunity memory over the population.

## II. METHODS

### A. Models of two consecutive strains

Let there be an outbreak of a first epidemic strain, Σ_1_, governed by SIR equations:

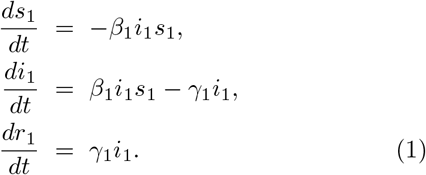

Here, *s*_1_(*t*), *i*_1_(*t*), and *r*_1_(*t*) are the fractions of susceptible, infected, and recovered individuals over time. They are normalized such that *s*_1_(*t*) + *i*_1_(*t*) + *r*_1_(*t*) = 1, thus only two of the equations above are needed to keep track of the epidemics. Occasionally, we will refer to non-susceptible individuals: *n*_1_(*t*) ≡*i*_1_(*t*) + *r*_1_(*t*). The parameters (*β*_1_, *γ*_1_) and the initial fractions of population in each compartment, 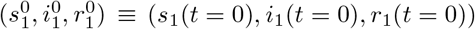, fully determine the unfolding of the outbreak.

We can extract some valuable information from these equations without solving them fully. For example, we can define the strain’s basic reproduction number:

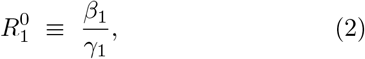

which corresponds to the spread of Σ_1_ in a fully susceptible population. We can also find out when the number of new infections stops growing:

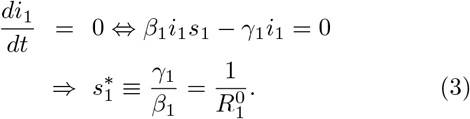

When the number of susceptible individuals is above the threshold defined by 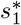, the infection grows. If the susceptible population is below this threshold, individuals already infected or recovered are so abundant that, in average, the virus cannot find new hosts to spread onto. Then, *herd immunity* has been reached and the outbreak remits spontaneously.

We can rewrite this condition as:

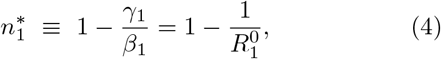

such that the outbreak starts remitting when 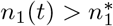. While 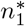 marks the peak of Σ_1_ infections, more incidental transmission is possible until it wanes off completely.

It is possible to compute the total population fraction, 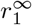, that will eventually undergo infection if the outbreak plays out naturally [25]:

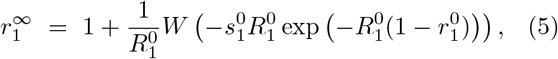

where *W* (*·*) is Lambert’s *W* function and 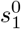 and 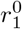 are initial conditions as introduced above (for 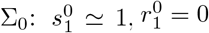).

Let us assume that, well after the first outbreak has played out, a second strain, Σ_2_, arrives which follows similar equations but with parameters (*β*_2_, *γ*_2_). This strain will be subjected to similar dynamics depending on whether the corresponding threshold 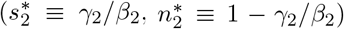 has been reached. How might cross-immunity provided by Σ_1_ alter this condition?

We introduce this effect in two different ways. The first one assumes that Σ_1_ provides *sterilizing immunity* with a probability *ϕ* (Fig. 1**a**). This means that, out of all the individuals who underwent infection with Σ_1_, a fraction *ϕ* of them cannot become infected by (and, accordingly, cannot propagate) Σ_2_. The second kind of cross-immunity (Fig. 1**b**) assumes that those who underwent infection with Σ_1_ can become infected by Σ_2_, but they do so at a reduced rate, *β*_2_*/σ* (with *σ >* 1). If they become infected, they are as contagious as usual. We refer to this as *attenuating immunity*.

**FIG. 1.**
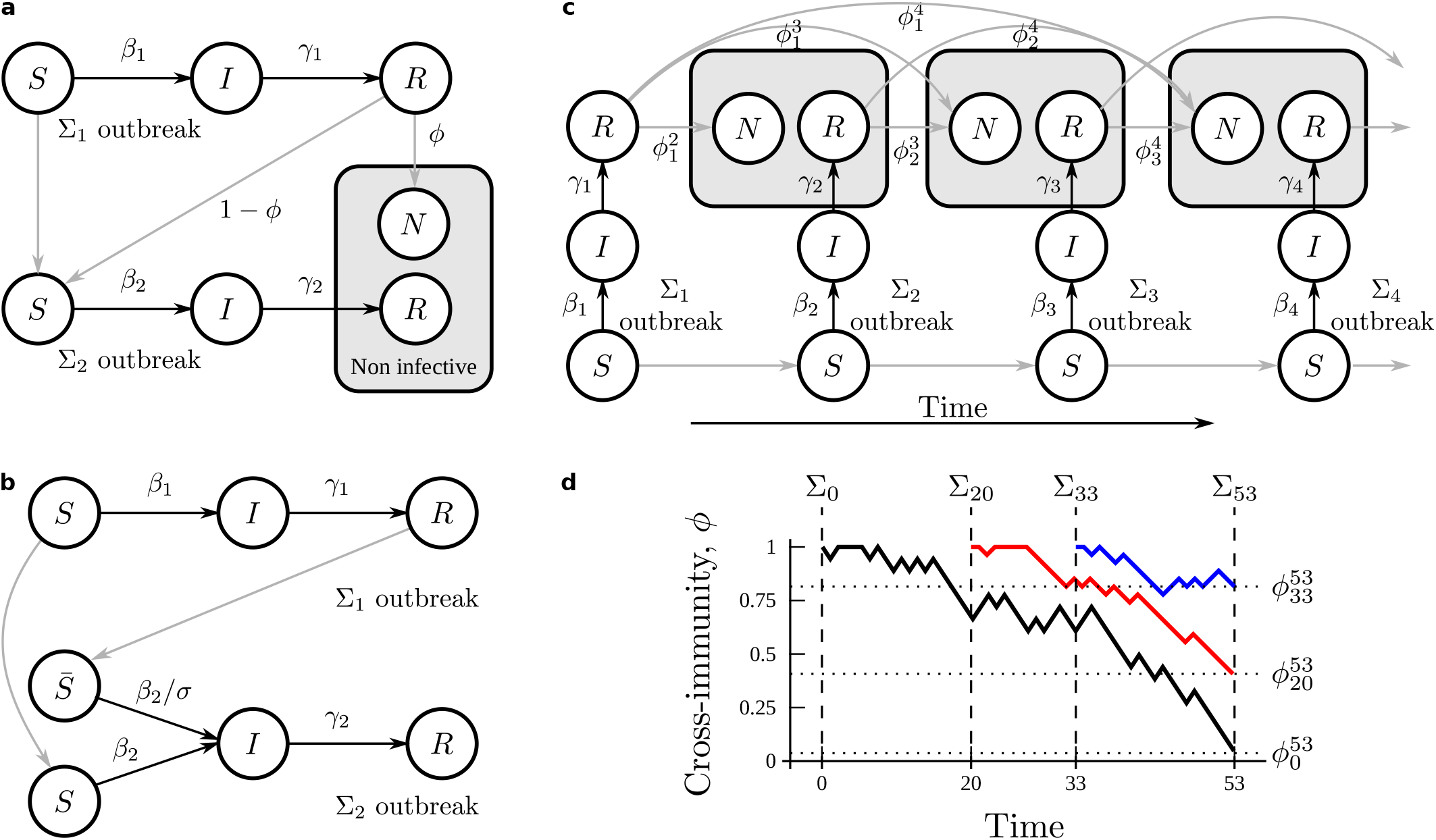
Seeking simplest models of cross-immunity. We depart from the SIR equations and attempt to modify them as little as possible to study cross-immunity between successive variants of a pathogen. **a** Model with *sterilizing immunity*. A strain Σ_1_ provides, with a probability 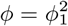, full cross-immunization against a strain Σ_2_. **b** Model with *attenuating immunity*. All individuals who underwent infection by Σ_1_ are partly protected against Σ_2_, such that their infection rate is reduced. **c** Rolling strains with sterilizing cross-immunity. Several variants of a same pathogen hit a population after each-other, each providing cross-immunity against future strains with probabilities 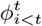. Immunity accumulates depending on rates of population renewal. **d** Levels of cross-immunity are modeled as a random walks. Two strains that arrive close in time are more likely to offer more protection against each-other. Variants that fail to cause an outbreak (e.g. because widespread sterilizing immunity prevents it) do not generate immunity responses against future strains.

We will use estimates of parameters from actual SARS-CoV-2 strains to illustrate relevant information that can be extracted from our model. Let us notate the original SARS-CoV-2 strain as Σ_0_ and some of its variants of concern as Σ_*α*_, Σ_*β*_, Σ_*δ*_, and Σ_*o*_ (with the subscripts carrying the standard names). We will use the same Greek-letter underscripts (and 0) to label model parameters when referring to SARS-CoV-2 strains, while we will use numbers 1 and 2 when referring to variants in the model.

### B. Model of rolling strains

To study longer-term effects of cross-immunity, we wish to learn how strains more distant in time might affect statistical features of outbreaks—such as their average size (measured as the population fraction they infect), typical SIR parameters of successful strains, or time between successive outbreaks of a given magnitude. We want to investigate how these properties are changed by the extent of immunizing memory across the population or by evolutionary mechanisms that might favor more dissimilar variants. We try to do so with the simplest model that builds upon insights from the previous section.

We will model *sterilizing immunity* only. To do so, we just need to keep track of individuals recovered from each strain—as will become clear later. To build a similar model for *attenuating immunity* we would need to keep track of individuals that might undergo infection with several strains and might acquire different levels of protection. Also, while we have a closed form for *r*^*∞*^ in the SIR model (Eq. 5, which will become important), we do not have a similar expression for the model that results from attenuated immunity.

Let us assume a series of strains, [Σ_1_, Σ_2_, …, Σ_*t*_, …], rolling one after another (Fig. 1**c**). At any time, *t*, there is only one active variant, Σ_*t*_, such that two outbreaks never overlap. We are hence assuming that the duration of an outbreak constitutes a unit of time. We also assume that these rolling strains are variants of a same pathogen—as it happens in COVID-19 waves. At a microscopic level, viral sequences mutate, resulting in new strains that descend from each-other with slight phenotype variations. We model this relatedness by generating SIR parameters of successive variants from an uncorrelated, unbiased random walk:

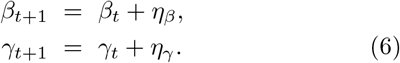

Here, *η*_*β*_ and *η*_*γ*_ are stochastic variables drawn from a Gaussian distribution with mean 0 and standard deviations *σ*_*β*_ and *σ*_*γ*_ respectively (in most simulations, *σ*_*β/γ*_ = 0.1). Both *β* and *γ* are positive, thus the random walk is bounded from below. We also set arbitrary upper bounds at *β*^+^ = 10, *γ*^+^ = 10 to prevent unrealistically high infectivity and recovery rates.

Eq. 5 allows us to compute 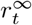 (the fraction of non-susceptible population after Σ_*t*_ completed its natural cycle). Since we consider rolling strains with cross-immunity, a part of this fraction (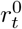, which we calculate below) consists of people immunized by earlier strains. This 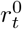works as an initial condition. Another part of 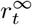 consists of people who properly underwent infection with Σ_*t*_. Let us note this later fraction of the total population as 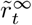. These are the people who might gain cross-immunity thanks to Σ_*t*_.

Let any earlier, successful strain, Σ_*i*_, provide cross-immunity against later variants, Σ_*j>i*_. This happens with a likelihood 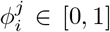 for each recovered individual. Thus, due to Σ_*i*_, a fraction 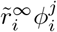 of the population is immunized against Σ_*j*_ when it hits. Similar fractions of cross-immunized population will be generated by other variants that preceded Σ_*j*_. These fractions might or might not overlap—i.e. a person might acquire sterilizing immunity against Σ_*j*_ from one, two, or more different ancestors. To calculate 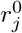, we need to compute the fraction of population who acquired cross-immunity from any (at least one) strain that preceded Σ_*j*_. Due to the possible overlaps, this becomes cumbersome. It is easier to note that the protected population is the complementary of the population who did not get immunity from any strain. This can be written as an equation more easily:

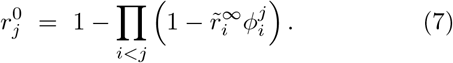

When Σ_*j*_ arrives, we can plug in this initial condition to the SIR equations to check the additional fraction of immunized population that Σ_*j*_ produces before it remits spontaneously:

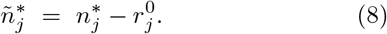

If 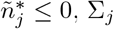 fails to spread in the population. If 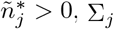 causes an outbreak. The amount of people infected by this new variant can again be calculated through Eq. 5 as 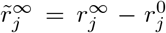. We identify 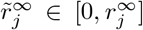 as the magnitude or size of the Σ_*j*_ outbreak.

For cross-immunity, relatedness between strains is important. We model it using random walks again, now bounded within 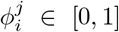 (Fig. 1**d**). Say that Σ_*i*_ produces an outbreak; but that the next strains, Σ_*i*+1_, …, Σ_*j−*1_, fail to spread; until Σ_*j*_ results in an outbreak again. We kick-start a random walk at *t* = *i* that begins with 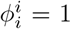 (i.e. Σ_*i*_ offers full protection against itself). For each successive strain, whether they produce an outbreak or not, we keep track:

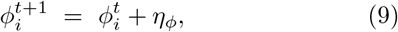

with *η*_*ϕ*_ a stochastic Gaussian variable of mean 0 and standard deviation *σ*_*ϕ*_ (usually *σ*_*ϕ*_ = 0.1). Non-spreading variants do not produce cross-immunity, hence we do not start random walks for any of them. After Σ_*j*_ unfolds, it does generate cross-immunity, and a new random walk is started with 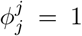. Protection against future strains might be gained from either Σ_*i*_ or Σ_*j*_ now, thus the former random walk remains active. Nothing forces these random walks to offer less cross-immunity over time, meaning that a strain might offer higher protection for variants further in the future than for closer ones. This is counter-intuitive, but it has happened for SARS-CoV-2 waves as Σ_0_ offered higher cross-immunity against Σ_*δ*_ than against Σ_*α*_ and Σ_*β*_ (Fig. 3) [24].

We should keep a random walk running for each successful strain, but carrying so many parallel processes is computationally expensive. To solve this problem, we introduce another relevant ingredient that is well grounded in biology and epidemic dynamics—population renewal. We assume that, at each time unit (i.e. after each new strain’s cycle), a fraction *δ* of the population is renewed. This has no effect for people who did not acquire immunity—as they are replaced by new people who are not immunized either. On the other hand, the population fraction that underwent infection by Σ_*i*_ becomes a time-dependent decaying variable:

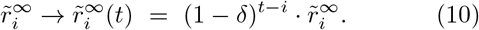

To ease the computational load, we stop tracking any variant whose 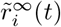 falls below 1% of population size.

The parameter *δ* introduces a measure of immune memory across the population. If *δ* = 1, no memory is ever left of successful strains. We obtain a trivial series of SIR outbreaks with parameters drawn from a random walk. For *δ <* 1, the population keeps a memory of the epidemics it endured—thus affecting which future strains can possibly succeed and, hence, which model parameters are likely to generate outbreaks.

## III. RESULTS

### A. Cross-immunity between two consecutive strains

#### 1. Sterilizing immunity

The mechanism for sterilizing cross-immunity leaves the equations for the second strain (Eqs. 1) unchanged, but alters their initial conditions—a population fraction 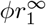 starts as non-susceptible (Fig. 1**a**). The derivative of infections at the second outbreak onset reads:

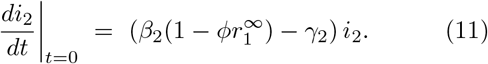

By equating this to zero, we find a threshold that determines whether sterilizing cross-immunity can prevent the outbreak:

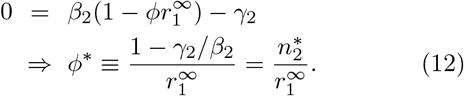

If *ϕ > ϕ*^*∗*^, Σ_1_ provides enough sterilizing cross-immunity. From the beginning, Σ_2_ cannot find sufficient hosts— any impending outbreak remits spontaneously. If *ϕ < ϕ*^*∗*^, enough susceptible individuals remain for Σ_2_ to take hold. Fig. 2**a** shows the amount of cross-immunity needed to avoid the second outbreak, *ϕ*^*∗*^, as a function of 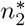 and 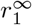. For 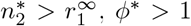; hence the spread of Σ_2_ becomes unavoidable.

**FIG. 2.**
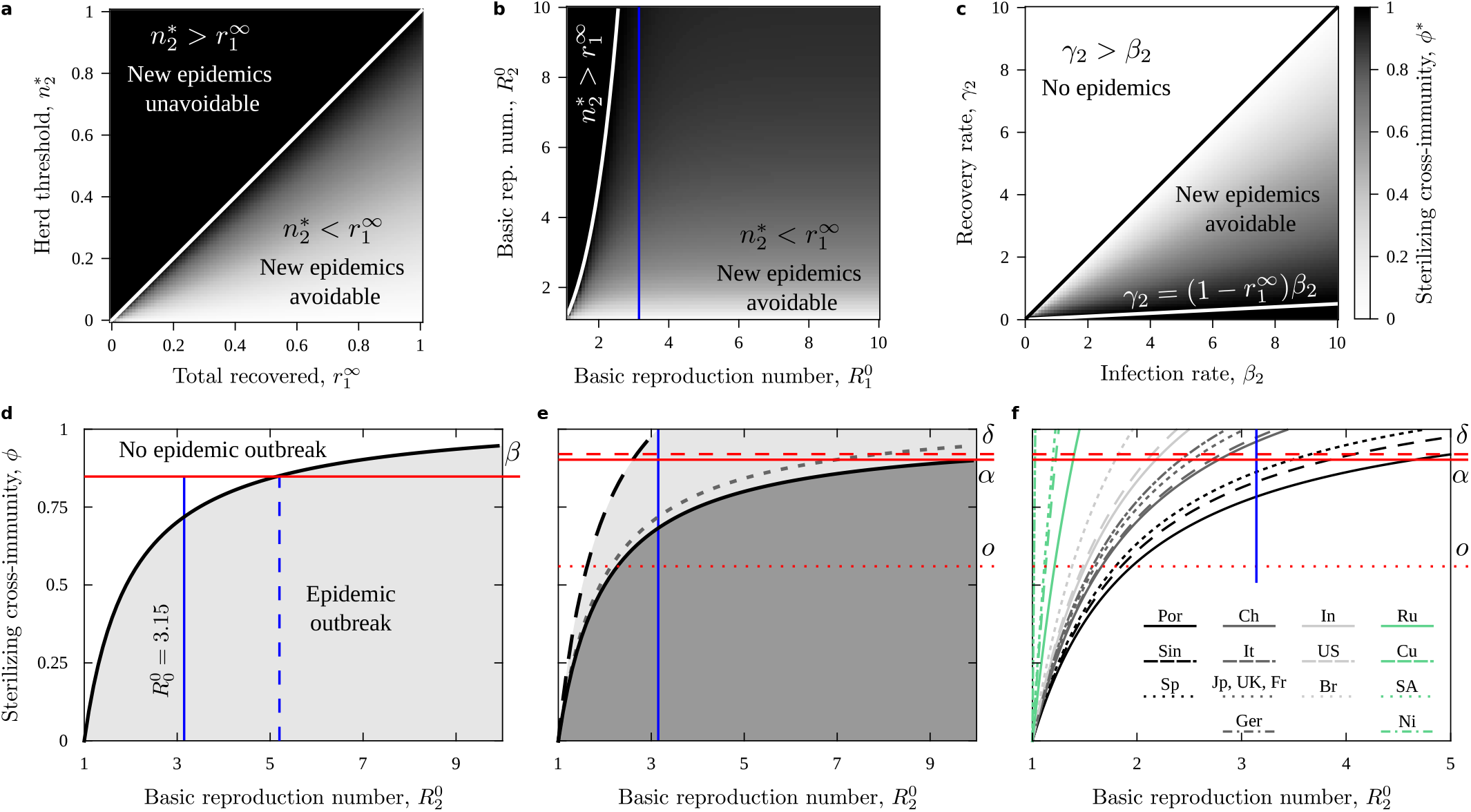
Sterilizing cross-immunity between two successive strains. **a** Minimum cross-immunity needed for Σ_1_ to prevent Σ_2_ outbreaks as a function of Σ_1_-recovered individuals, 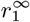, and Σ_2_-herd-immunity threshold,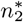. The white line marks when outbreaks become unavoidable (Σ_1_ cannot generate enough protection). **b** Same, as a function of basic reproduction numbers, 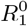 and 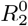. The white curve (mapped non-linearly into this representation) again bounds unavoidable outbreaks. A vertical blue line marks *R*^0^ = 3.15, a robust estimate of *R*^0^ for the original SARS-CoV-2 virus [22, 23]. **c** Same, as a function of Σ_2_ model parameters, *β*2 and *γ*2; assuming 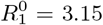. Unavoidable outbreaks appear confined to a small region (lower-right corner). **d-f** Cross-immunity in SARS-CoV-2. **d** Protection if the original virus, Σ_0_, had played out its natural SIR cycle. The black curve (cross-section of panel **b** with *R*^0^ = 3.15) separates all variants that would have been avoided in this scenario (white region). The horizontal red line marks an estimated level of Σ_0_-elicited protection against Σ_*β*_ [24]. If Σ_0_ had played out naturally before Σ_*β*_ hit, Σ_*β*_ outbreaks would have been halted unless 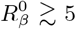. **e** Protection assuming 100% vaccination rate (solid black curve), and assuming ‘just enough’ Σ_0_ infections before its remission (i.e. only 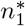 people got infected, dashed black curve). Protection as in **d** (dashed gray curve) is shown as reference. Red lines mark Σ_0_-elicited cross-immunity estimated against Σ_*α*_ (solid), Σ_*δ*_ (dashed), and Σ_*o*_ (dotted). Both Σ_*α*_ and the deadly Σ_*δ*_ would have been prevented in fully vaccinated populations unless they had and impressive *R*^0^ ≳ 10. **f** Protection offered by vaccination rates (2 doses) in Portugal (Por), Singapore (Sin), Spain (Sp), China (Ch), Italy (It), Japan (Jp), United Kingdom (UK), France (Fr), Germany (Ger), India (In), United States (US), Brazil (Br), Russia (Ru), Cuba (Cu), South Africa (SA), and Nigeria (Ni) as of October 16, 2021. Note the scale in the horizontal axis (different from **d, e**). Estimated cross-immunity from Σ_0_ is marked (red lines), but estimates of vaccine efficiency could be used instead.

*ϕ*^*∗*^ only depends on model parameters through the basic reproduction numbers 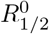. This makes it easier to use empirical estimates from real outbreaks. Fig. 2**b** shows *ϕ*^*∗*^ as a function of 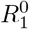 and 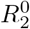. The condition for unavoidable outbreaks 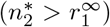 becomes non-linear in this representation. A vertical blue line marks 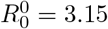, a robust estimate for the original variant, Σ_0_, of the SARS-CoV-2 virus [22, 23]. This allows us to visualize *ϕ*^*∗*^ as a function of infectivity, *β*_2_, and recovery rates, *γ*_2_, of potential variants for this actual virus (Fig. 2**c**).

The line marking 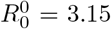 (Fig. 2**b**) does not intersect 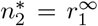 within the shown 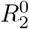 range. This means that all SARS-CoV-2 variants with 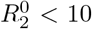 could have been blocked if Σ_0_ had played out completely *and* it offered enough sterilizing cross-immunity. This becomes more clear in Fig. 2**d** (which plots the relevant cross-section of Fig. 2**b**). For each possible new variant with 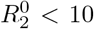, and assuming Σ_0_ played out fully, infecting a population fraction 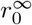, it exists a feasible level of sterilizing cross-immunity, 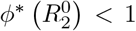 (black curve), that would prevent any new outbreaks.

In Fig. 2**d**, a horizontal red line marks the estimated protection that Σ_0_ offers against Σ_*β*_ [24]. (Actually, the likelihood that Σ_0_ infection prevented symptomatic Σ_*β*_ infection—hence a best-case scenario.) The intersection of this protection level with 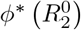 (dashed blue line) indicates all the variants that Σ_0_ would have protected against if it had played out naturally (infecting a population fraction 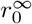). Roughly, any variant with 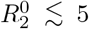 would have been prevented with this level of cross-immunity.

We can use similar plots to evaluate whether new variants should worry us given levels of cross-immunity elicited by a vaccine and fractions of vaccinated population. In Figs. 2**e, f**, horizontal red lines mark sterilizing protection (estimated as above [24]) provided by Σ_0_ against variants Σ_*α*_ (solid), Σ_*δ*_ (dashed), and Σ_*o*_ (dotted). We assume that vaccines prompt a same cross-immune response. The fraction of vaccinated population enters Eq. 12 as an initial condition, instead of 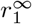, resulting in altered 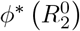 curves.

Fig. 2**e** shows two extreme cases: (i) just enough people to stop Σ_1_ was vaccinated (dashed curve) and (ii) the whole population was vaccinated (solid curve). Fig. 2**f** shows 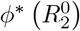 depending on vaccination rates across the world as of Oct. 16, before the deadly Σ_*δ*_ wave hit most of the represented countries. Estimates of 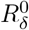 range between 3.2 and 6 [26–28]. In this best-case scenario, Σ_*δ*_ could have been avoided by fully vaccinated populations. Cross-immunity offered for Σ_*o*_ would not have been enough to stop it unless its basic reproduction number were very low (which, likely, was not [29, 30]).

#### 2. Attenuating immunity

Our second mechanism is closer to implementations of cross-immunity in the literature [5, 19–21]. Individuals who underwent Σ_1_ infection remain susceptible during a Σ_2_ outbreak, but their infection rate is attenuated: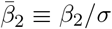, with *σ >* 1. This entails both a change of initial conditions and of model equations for Σ_2_ outbreaks (Fig. 1**b**):

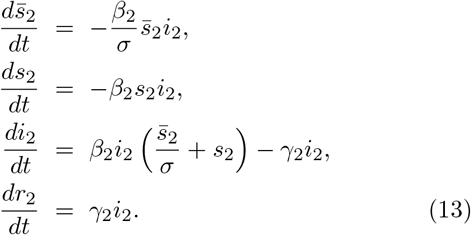

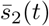 represents the fraction of susceptible individuals with attenuated infection rate, while *s*_2_(*t*) are regular susceptible hosts. Population is normalized again, making one of these equations redundant.

We check when Σ_2_ stops spreading:

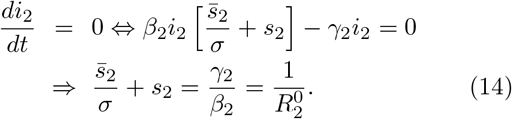

We get a herd-immunity threshold for the weighted sum of susceptible individuals: If 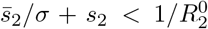, then Σ_2_ cannot cause enough new infections in average—the outbreak remits spontaneously. If 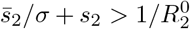, the outbreak grows until the susceptible populations become depleted enough. Both 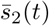 and *s*_2_(*t*) decrease over time always, thus they can only pass this threshold once—i.e. these dynamics cannot bounce back up.

We substitute the initial conditions after Σ_1_ to check whether Σ_2_ is spontaneously halted at its onset. A population fraction 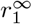 starts in 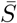, while 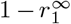 start out as regular susceptible individuals:

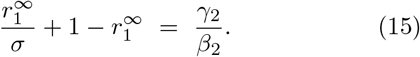

By solving for *σ*, we find a threshold that determines whether attenuating cross-immunity can prevent the outbreak:

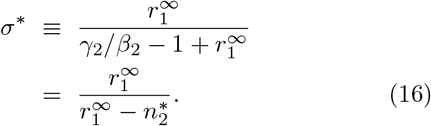

If *σ* is larger than this threshold, then the level of attenuation is enough as to halt a Σ_2_ outbreak.

For 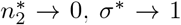 ; thus any attenuation is enough to halt Σ_2_. To reach this limit, *β*_2_ itself must become very small—hence explaining why any *σ >* 1 would suffice. On the opposite end 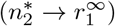, attenuating cross-immunity cannot be achieved since *σ*^*∗*^. Eq. 16 has a singularity at 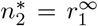, after which *σ*^*∗*^ becomes negative (Fig. 3**a**), and is thus meaningless for 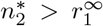. But we know that, due to parsimony, there is no level of attenuation that can prevent outbreaks in that regime. Thus, we recover the same limit condition as for sterilizing cross-immunity. These results, again, only depend on model parameters through the basic reproduction numbers. Fig. 3**b** shows *σ*^*∗*^ in the 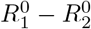 plane. We mark the boundary of unavoidable outbreaks (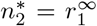, red) and the estimate 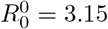 for the original SARS-CoV-2 strain (blue). By holding this value fixed, we can represent 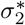 as a function of *γ*_2_ and *β*_2_ (Fig. 3**c**).

**FIG. 3.**
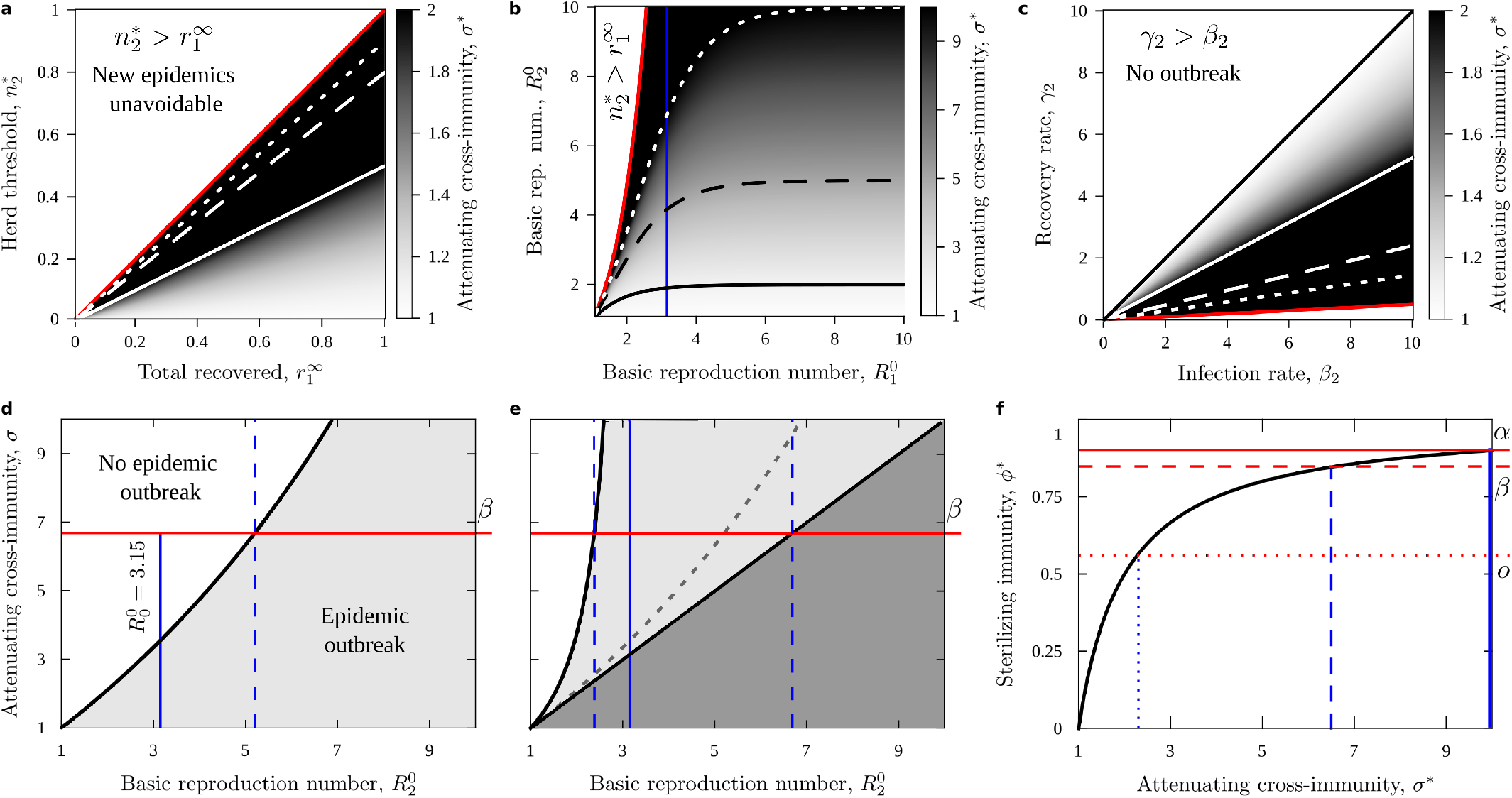
Effects of attenuating cross-immunity. **a-c** Same as in 2**a-c**. The cross-immunity parameter now spans *σ∈* [1, *∞*), and has been cropped to aid the visualization (note differences in color bars, which were adjusted to enhance the displayed gradients). Solid (black or white) curves indicate *σ* = 2. Dashed (black or white) curves indicate *σ* = 5. Dotted white curves indicate *σ* = 10. Red curves indicate the boundary beyond which no level of cross-immunity can avoid new outbreaks. These regions match those for sterilizing immunity in all parameter spaces, anticipating a mapping between both cross-immunity mechanisms. **d, e** Same as in 2**d, e. d** The dashed vertical line marks the level of protection achieved by the original SARS-CoV-2 virus against the *β* variant. To achieve the same level of protection with attenuating immunity, we seek the intersection of this vertical line with the black curve (which is, in turn, a vertical cut of panel **b** with *R*^0^ = 3.15). **f** Eq. 17 (black curve) formalizes the equivalence between both models. Levels of attenuating immunity needed to match estimated sterilizing immunity of real SARS-CoV-2 variants can be read from the intersections of horizontal red lines and the corresponding vertical marks.

Fig. 3**d** plots the cross-section of Fig. 3**b** with 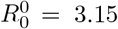. The black curve marks 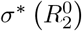, the attenuation level needed to halt new SARS-CoV-2 strains as a function of 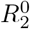. The dashed blue line marks the separation between strains that could and could not be halted by sterilizing cross-immunity. The red horizontal line uses 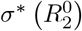 to track back the level of attenuation that would offer a similar protection: Σ_2_ infection following Σ_0_ recovery should be ∼ 7 times less frequent than natural infection. Similar *translations* are possible for scenarios with population immunized through vaccination (Fig. 3**e**).

We make this equivalence between models rigorous by demanding that the 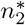 featured in Eqs. 12 and 16 take the same value, obtaining:

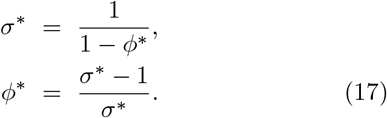

Fig. 3**f** shows this mapping between models. We mark levels of attenuating immunity needed to match sterilizing immunity for SARS-CoV-2 variants. Eq. 17 does not depend on the fraction of immunized population— suggesting a deep correspondence between both immunizing mechanisms.

### B. Cross-immunity between rolling strains

Model dynamics alternate (i) long periods of no activity with periods of (ii) sparse and (iii) very frequent outbreaks. The former happen when the random walk is confined to the upper-left half of the (*β, γ*) plane (no-epidemics region, Fig. 2**c**). We name this area *Z*_0_, and we call *Z*_1_ to the lower-right hemiplane (where SIR outbreaks always happen). If the random walk spends enough time in *Z*_0_, all memory about earlier outbreaks is lost. Fig. 4**a1** shows model behavior during sparse outbreaks. Such sparsity might be due to the random walk dwelling near the *Z*_0_ *− Z*_1_ boundary. Decay dynamics following population renewal stand out. The fraction of protected population remains similar to outbreak magnitudes. Fig. 4**a2** shows model behavior during frequent outbreaks. This regime corresponds to deep dives into *Z*_1_. New successful strains appear at every time step. The dynamics are dominated by the accumulated immunized population. The fraction of protected population is much larger than the average outbreak size.

**FIG. 4.**
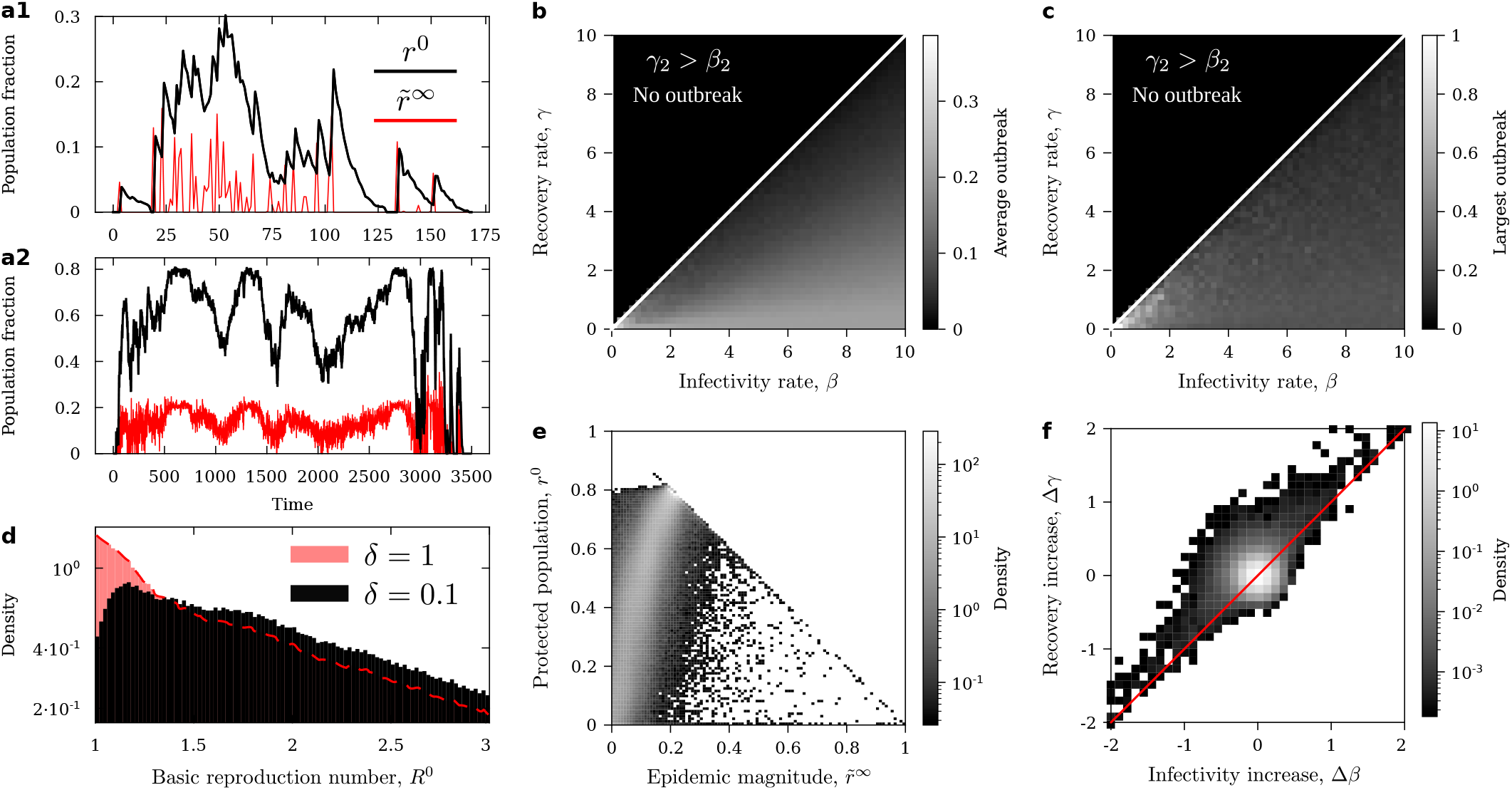
Model of rolling strains with fixed population renewal, *δ* = 0.1. **a** Example of the model dynamics during sparse (**a1**) and frequent (**a2**) epidemic outbreaks. We show the population fraction immune to each new strain (*r*^0^, black), and each outbreak’s magnitude (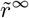,red). **b** Average outbreak magnitude as a function of model parameters *β* and *γ*. **c** Largest outbreak as a function of model parameters *β* and *γ*. **d** Probability density function of the basic reproduction number among successful strains with cross-immunity (black) compared to successful strains in the standard SIR model (red). A red dashed curve outlines the distribution of SIR reproduction numbers to show that both scenarios show similar tails for large *R*^0^. **e** Joint probability density function of epidemic magnitudes, 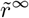, and protected fraction of the population when each successful strain hits, *r*^0^. **f** Probability density function of increases in model parameter between two consecutive successful strains.

We are interested in statistical properties of successful strains over long periods. Therefore we ran our model for 10^6^ time steps under different experimental conditions.

Fig. 4**b** shows average outbreak magnitude as a function of model parameters (*β, γ*) with *δ* = 0.1. Outbreak sizes are greatly reduced with respect to SIR processes without cross-immunity (Sup. Fig. 1**f**). Fig. 5**a** shows that outbreak size mitigation depends linearly on population renewal. Both with (Fig. 4**b**) and without (Sup. Fig. 1**f**) cross-immunity, outbreak magnitude becomes larger the deeper we dive into *Z*_1_. But only with cross-immunity, relatively large outbreaks stand out around the lower-left corner of parameter space, near the *Z*_0_*− Z*_1_ boundary. This becomes more evident when plotting the largest outbreak as a function of model parameters (Fig. 4**c**). With cross-immunity, the largest outbreaks happen exclusively in this small region, while larger outbreaks happen for broader combinations of parameters without cross-immunity (Sup. Fig. 2**f**). Indeed, if the random walk reaches deep into *Z*_1_, it will follow a long path with frequent successful strains, thus accumulating immunized population unless renewal is high.

**FIG. 5.**
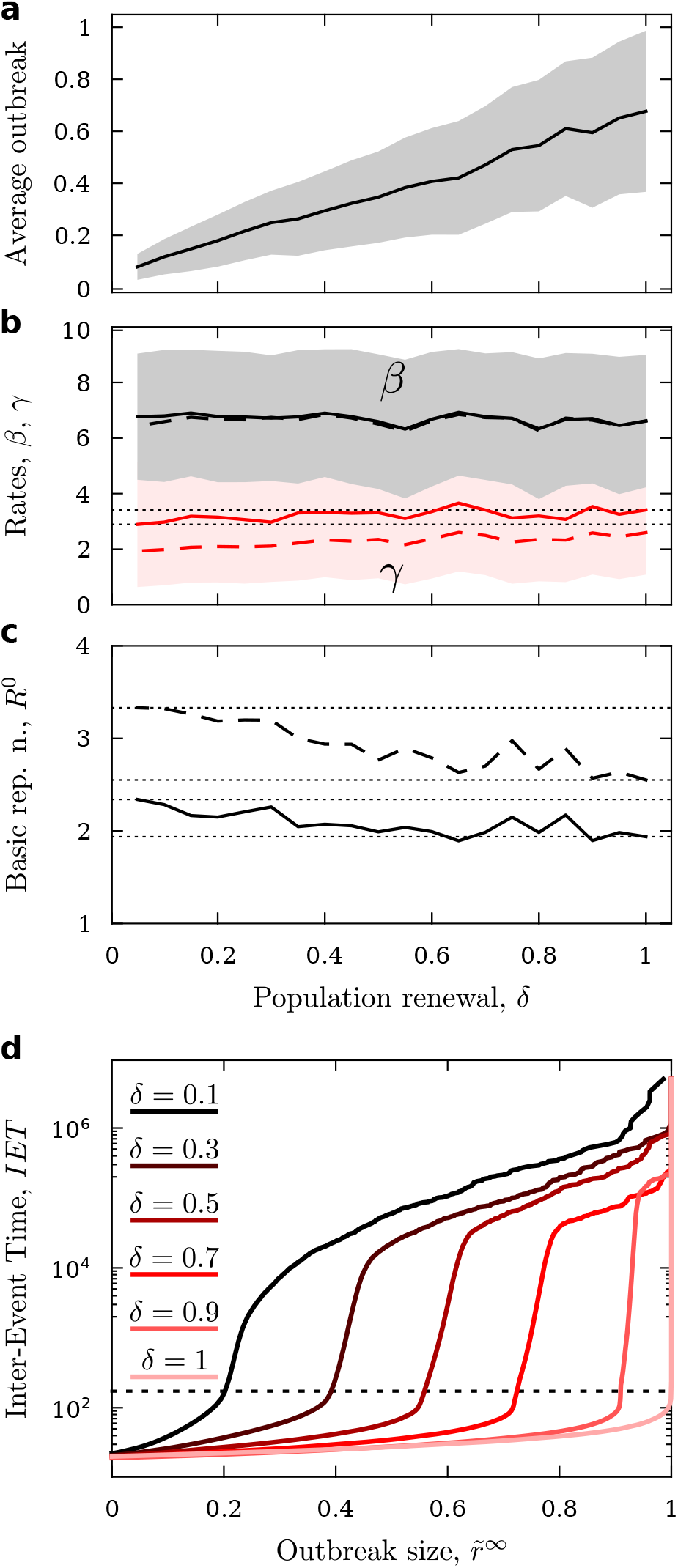
Model of rolling strains—comparison with varying population renewal. **a** Average outbreak magnitude as a function of population renewal. **b** Average model parameters *β* (black) and *γ* (red) of successful strains as a function of population renewal. We observe a striking constancy. Solid curves represent unweighted averages and shading represents unweighted standard deviations. Dashed curves represent averages weighted by outbreak size. Dotted horizontal lines help visualize the slight increase in *γ* from its value at *δ* = 0.05 to *δ* = 1. **c** Basic reproduction numbers from average model parameters from panel **b** (both unweighted, solid, and weighted, dashed) as a function of population renewal. Dotted horizontal lines help visualize trends within the studied range. **d** Inter-event interval as a function of event size for several values of population renewal.

Notwithstanding this localization of large outbreaks for small renewal (hence large cross-immunity), the average *β* and *γ* appear relatively constant for varying *δ* (Fig. 5**b**). This suggests that, overall, cross-immunity prevents (or weakens) outbreaks similarly throughout parameter space. Upon close inspection, these averages actually change slightly. Dotted lines in Fig. 5**b** help visualize that *γ*(*δ* = 0.1) *< γ*(*δ* = 1). This effect becomes more salient in the basic reproduction number (Fig. 5**c**, since *R*^0^ = *β/γ* has *γ* in the denominator, hence amplifying trends). *R*^0^ stands for a growth factor, thus its variation has an exponential effect in the dynamics. Changes in this parameter with respect to SIR without cross-immunity stand out in distributions of *R*^0^ for successful strains (Fig. 4**d**). Distributions for both *δ* = 0.1 (black) and *δ* = 1 (red) present similar tails, but their peaks (at 1 and *∼* 1.2 respectively) differ.

A population’s current immunization state constrains the characteristics of the next, possible successful strains. Fig. 4**e** shows how a given amount of protected population limits the magnitude of possible outbreaks. This effect wanes as increased population renewal erases the immune memory (Sup. Fig. 3). A current immunization state also influences likely increases in model parameters, *β* and *γ*. The underlying random-walk is unbiased, but the distribution of *β* and *γ* increases becomes asymmetric (Fig. 4**f**). If a random-walk step increases *β* and decreases *γ* (deeper into *Z*_1_), the strain becomes more aggressive, likely producing an outbreak despite existing cross-immunity. If the step goes in the opposite direction (towards *Z*_0_), the resulting, less-aggressive strain might be averted by population immunity. Then, the next outbreak shall arrive when memory is lost and the random walk has drifted further away. This asymmetry is lost as population renewal increases (Sup. Fig. 4). Fig. 4**f** also shows frequent events along the diagonal Δ*β* = Δ*γ*. These events result from returns from *Z*_0_ into *Z*_1_ (reentry points can be arbitrarily far apart along the *Z*_0_*− Z*_1_ boundary).

Average time between consecutive outbreaks of a given size (Inter-Event Times, *IET*) grows monotonously and include a stark non-linearity for any *δ* ≠ 1 (Fig. 5**d**). The step-like behavior suggests a singular, *δ*-dependent outbreak size, *r*^*∗*^(*δ*), such that larger events very swiftly become very rare (roughly, at the intersection of *IET* curves with the dashed black line, Fig. 5**d**). This invites a qualitative distinction between epidemic (frequent, small magnitude) and pandemic (rare, larger size) dynamics that emerges naturally out of cross-immunity (note that *r*^*∗*^(*δ* = 1) = 1, thus events of all sizes are fairly equally frequent in memory-less SIR).

## IV. DISCUSSION

We studied simple models of cross-immunity to see how it affects dynamics in short- and long-term scopes. Starting from the SIR equations (a cornerstone of epidemic dynamics), we added as little as possible to include cross-immunity, avoiding additional dynamic ingredients (e.g. reinfection, strain coexistence and competition, population structure, etc.) present in other cross-immunity models [5, 19–21].

Regarding short-time effects, we investigated two mechanisms: (i) A past strain *might* provide *sterilizing immunity* against a future strain. Each recovered individual has a chance of becoming sterile to the second pathogen. (ii) A past strain *does* cause *attenuating immunity* against a future strain. All affected individuals might become infected by the second variant, but at a lower rate. Both mechanisms modify the thresholds of herd immunity of the standard SIR model. Our thresholds depend on the magnitude of the first outbreak, and on the likelihood of sterilization (in the first case) or on the level of attenuation (in the second).

These thresholds result in intuitive maps that separate which future strains can and cannot cause outbreaks. These plots also asses whether vaccination rates and vaccine efficiency suffice to stop new variants—a point illustrated with actual data from SARS-CoV-2 waves. The reliability of these charts depends on accurate estimates of vaccine efficiency and each variant’s basic reproductive number. Measuring these parameters might require that a strain spreads throughout a population (which would preempt the utility of our approach). However, lab experiments can compare emerging variants to earlier ones, rendering quick proxies to build preliminary maps for variants of concern.

Discussion regarding herd protection usually considers sterilizing immunity alone. Hopes of herd immunity against SARS-CoV-2 often faltered as it became evident that vaccines would not be sterilizing [7]. Following the same logic, several research strategies bet on sterilizing vaccines as a more definitive way out of this and other pandemics [31–35]. Our results show that attenuating immunity also modify SIR thresholds substantially. We further prove an equivalence (encapsulated by Eqs. 17) between both immunity mechanisms, offering new hope in dealing with epidemic scenarios.

In a relevant perspective, Lipsitch et al. [7] reviewed possible epidemiological scenarios for the COVID-19 pandemic depending on cross-reactive activity developed by different kinds of T-cells localized in distinct tissues. Three cases are studied: (i) reduction of lung burden, in which antibodies only mitigate the severity of late-stage COVID disease; (ii) accelerated antibody response, which mitigates the disease and shortens its infectivity period; and (iii) fast and strong response in the upper respiratory tract (URT), which would halt within-organism spread swiftly, and which comes the closest to sterilizing immunity. Implications to the epidemiological dynamics are discussed for all cases, but only in the third one (which the authors consider the less likely) do they expect a reduced herd-immunity threshold.

We should revise all three scenarios in a more optimistic note. On the one hand, even if it is unlikely that URT response is prompt and strong enough as to be sterilizing, it might still result in a reduced infectivity rate. Hence, even a weak response of the third kind could trigger our mechanism of attenuating cross-immunity. On the other hand, both (i) and (ii) cases result in reduced viral load and duration of the infective process, lowering the chances that cross-immunized individuals infect others. This effect is different from the one that we model, but it too results in a reduced infectivity (like *β/σ*) for a population subset. Based on our results, we expect changes in the herd-immunity thresholds in all scenarios. Herd immunity is a stark non-linear effect that causes spontaneous remission of epidemic outbreaks. Of all epidemiological consequences of cross-immunity discussed in [7], more lenient thresholds is the most impactful and desired possibility.

Regarding long-time effects, we simulated a succession of strains rolling one after the other and offering cross-immunity over longer periods of time. The length of these periods is loosely controlled by population renewal—the smaller it is, the larger the population-wide effect of cross-immunity.

Increased cross-immunity reduces the average epidemic magnitude linearly. When population renewal is small, the largest outbreaks become confined to a small region of parameter space—while, in memory-less SIR processes, large outbreaks happen for broad combinations of infectivity and recovery rates. Thus, cross-immunity alters the phenotype of strains from which extreme events can be expected. In our model, a natural distinction appears between frequent, relatively small outbreaks and rare, relatively large episodes. This distinction (not present in SIR processes) invites an emergent classification of “epidemic” and “pandemic” events.

Existing cross-immunity levels constrain which model parameters (hence which strain phenotypes) might produce an outbreak next. Our model (built upon an unbiased random walk) has no explicit preference regarding whether infectivity and recovery rates should increase or decrease as strains roll out. Cross-immunity induces an asymmetry favoring more infective strains with longer recovery rates.

There is ample room for future exploration regarding this last point. We do not distinguish between recovered and deceased individuals, nor do we track disease severity. Very likely, pathogens causing more serious illness, with increased death tolls, will induce stronger (cross-)immunity. This should further change where, in parameter space, large outbreaks are expected; as well as the phenotypes into which an active pathogen (e.g., currently, SARS-CoV-2) might evolve as constrained by cross-immunity. We modeled one of these aspects using random walks biased towards less averted strains (i.e., lower *ϕ*; not shown). We did not find qualitative differences from the behavior reported in this paper. This indicates that more ingredients are needed to expand the model in this direction.

A limitation of our model with rolling strains is that we only implement sterilizing immunity, as attenuating immunity poses some computational challenges. However, we can use the equivalence captured by Eq. 17 to find out what strains would have had similar effects using our second mechanism.

A more general limitation is in assuming SIR equations throughout. More factors, both biological (incubation period, super-spreaders, etc.) and social (confinement, fear, social distancing, etc.), affect the empirical parameters from SARS-CoV-2 analyzed. Our charts should be taken as limit cases. It is noteworthy, however, that we can produce meaningful tools for a real, ongoing epidemic process—this should be furthered. Note also that most of these factors would slow viral spread compared to actual SIR processes. Thus, more accurate models should result in more lenient thresholds.

The SARS-CoV-2 pandemic made explicit a very important limitation of SIR-like models: while they reveal overall trends and the existence of phenomena such as herd-immunity thresholds, they are useless to predict even the most salient aspects of a specific outbreak (namely, its magnitude and duration) [10]. We explore our models with the same philosophy, avoiding specific predictions and seeking broad qualitative changes induced by cross-immunity. To overcome the problems of compartmental models, and allow us to apply them in more realistic situations, two strategies are being explored: (i) To produce more detailed equations, that include as many aspects as possible. In this approach, cross-immunity should be indispensable, as the interactions between SARS-CoV-2 variants illustrate. (ii) To gather data at a microscopic level, registering sub-regional incidences, capturing population movements across road or airplane networks [14, 18, 36, 37], or quantifying the structure of social contact networks (often depending on age and cultural aspects of a given group) [38]. Regarding prediction, these details might work similarly to how abundant, localized data alleviates the chaotic weather dynamics—but gathering such low-level data in human behavior has a distinct ethic dimension that makes it problematic. Regarding qualitative aspects of epidemic dynamics, such microscopic data might help us understand where (in a spatial or social network) (cross-)immunity could be more effective— and hence design, e.g., smart quarantine or vaccination strategies [17, 36].

## Data Availability

All relevant data is contained or can be reproduced from the equations in the manuscript.

## Acknowledgments

We are indebted to Susanna Manrubia for her support during this research, and for the lively discussions that guided and improved our work. LFS wishes to acknowledge Jorge Mira, Alberto P Muñuzuri, Iván Area, Juan J Nieto, Pablo Boullosa, Adrián Garea, and Alejandro Carballosa for the initial discussions that fostered this work.

## Funding

Both IA-D and LFS were supported by grant PID2020-113284GB-C21, funded by MCIN/AEI/10.13039/501100011033. LFS has received funding from the Spanish National Research Council (CSIC) and the Spanish Department for Science and Innovation (MICINN) through a Juan de la Cierva Fellowship (IJC2018-036694-I), and from the Spanish “Instituto de Salud Carlos III and the Ministerio de Ciencia e Innovación” through the research grant COV20/00617 entitled PREDICO. The Spanish MICINN has also funded the “Severo Ochoa” Centers of Excellence distinction for the CNB, where this research was carried out (grant SEV 2017-0712), and the special grant PIE 2020-20E079 (awarded to the CNB) entitled “Development of protection strategies against SARS-CoV-2”.

## Author contribution

LFS conceived the research. IA-D and LFS performed calculations, carried out simulations, elaborated figures, and wrote the manuscript.

---

**SUP. FIG. 1.**
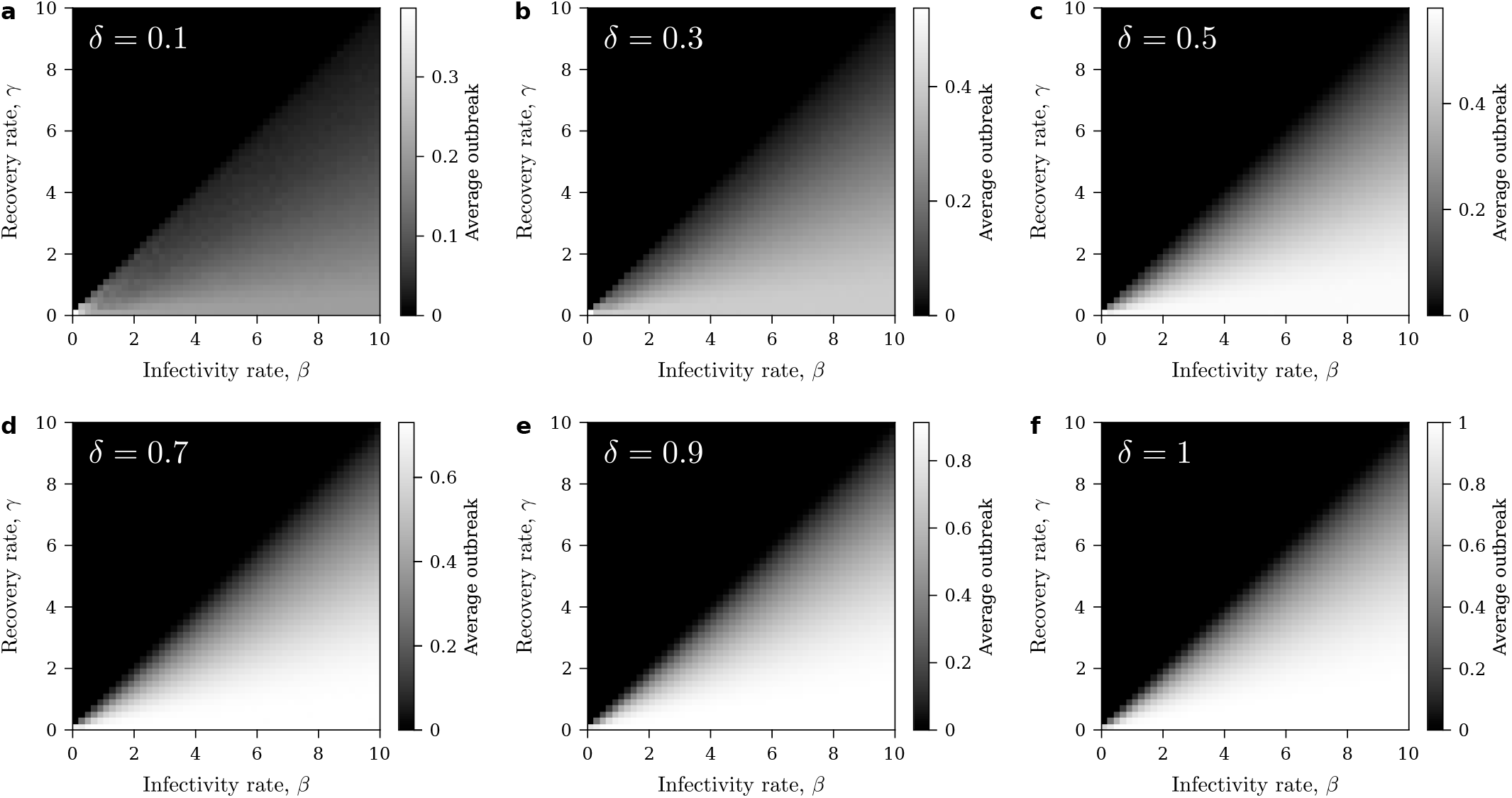
Average outbreak size as a function of model parameters in the rolling strings model. Death rates, **a** *δ* = 0.1, **b** 0.3, **c** 0.5, **d** 0.7, **e** 0.9, **f** 1.As δ → 1, a random sampling of SIR model outbreaks is recovered.

**SUP. FIG. 2.**
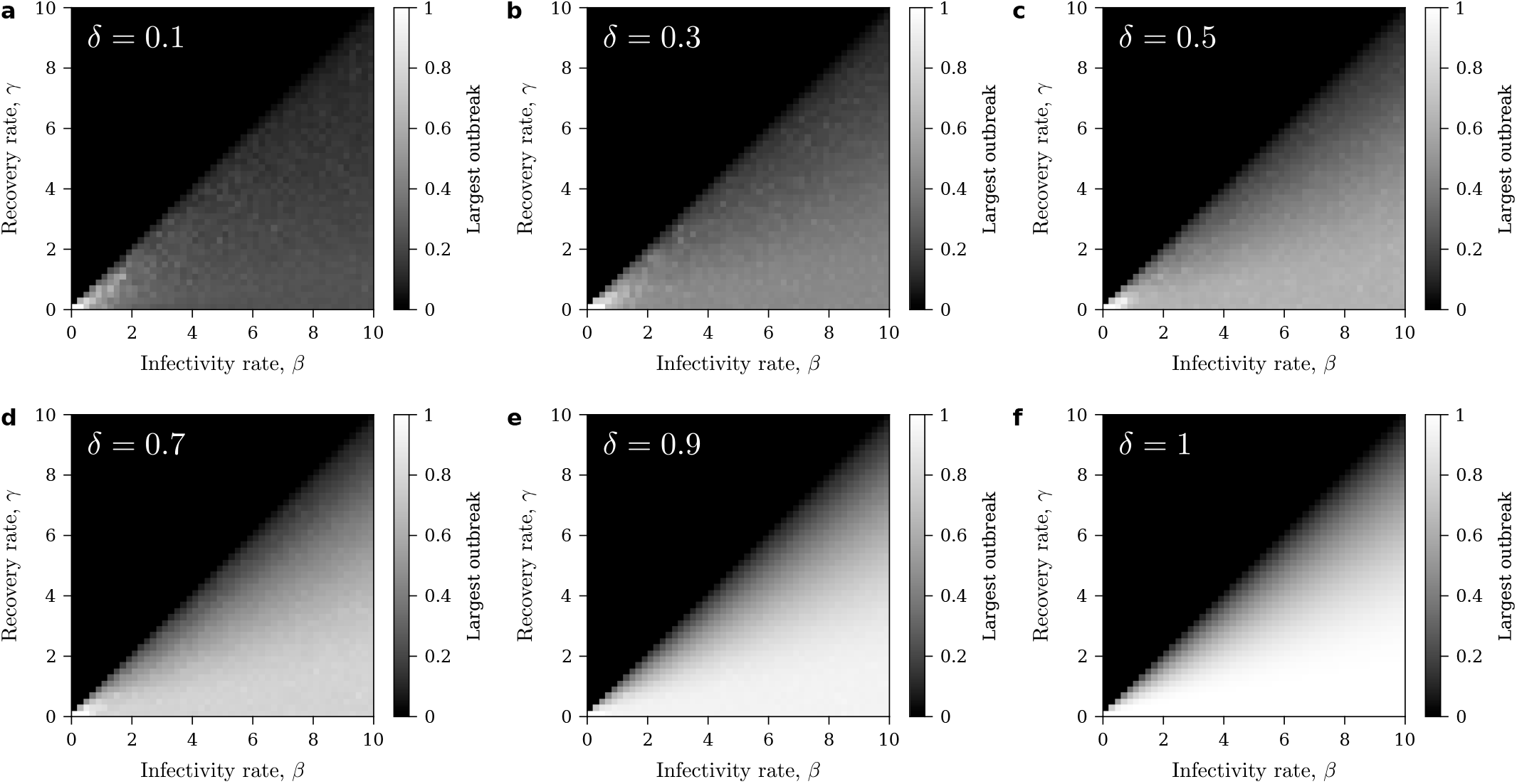
Largest outbreak size as a function of model parameters in the rolling strings model. Death rates, **a** *δ* = 0.1, **b** 0.3, **c** 0.5, **d** 0.7, **e** 0.9, **f** 1. For small enough population renewal (i.e. large cross-immunity memory), the largest outbreaks are confined to a small region of parameter space (bottom left, near the *Z*_0_ − *Z*_1_ border). When cross-immunity is removed (as *δ →* 1), very large outbreaks happen for almost any combination of model parameters.

**SUP. FIG. 3.**
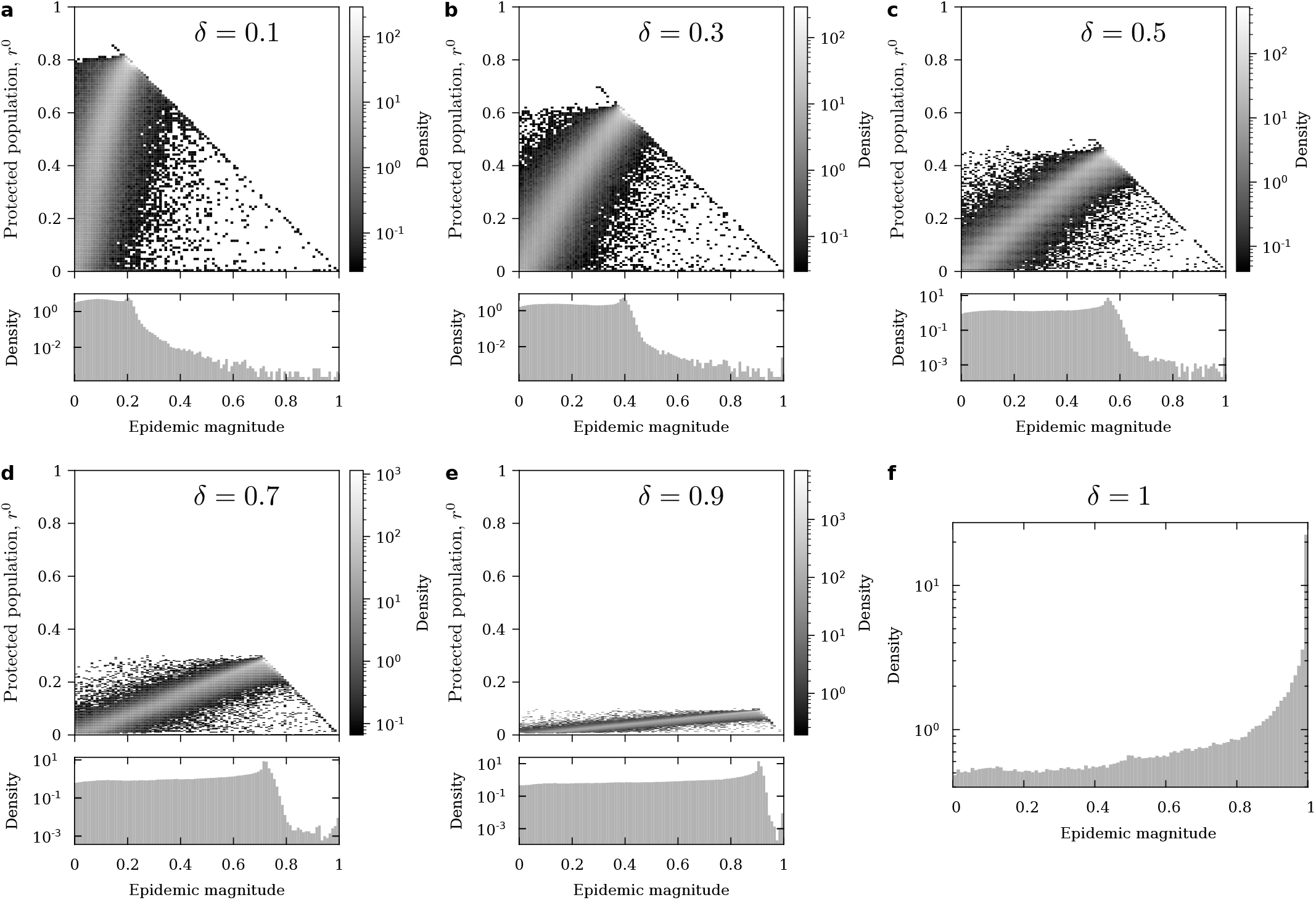
Joint probability density distribution of outbreak size and fraction of protected population in the rolling strings model. Death rates, **a** *δ* = 0.1, **b** 0.3, **c** 0.5, **d** 0.7, **e** 0.9, **f** 1. Note that, immediately after an outbreak of size *r*, at most an outbreak of size 1 − (1 − *δ*)*r* is possible. Note also that there is no protected population for *δ* = 1, thus the 2 − *D* probability density distribution reduces to a 1 − *D* histogram. The corresponding probability density distribution of outbreak size alone is displayed in all cases (bottom of each panel).

**SUP. FIG. 4.**
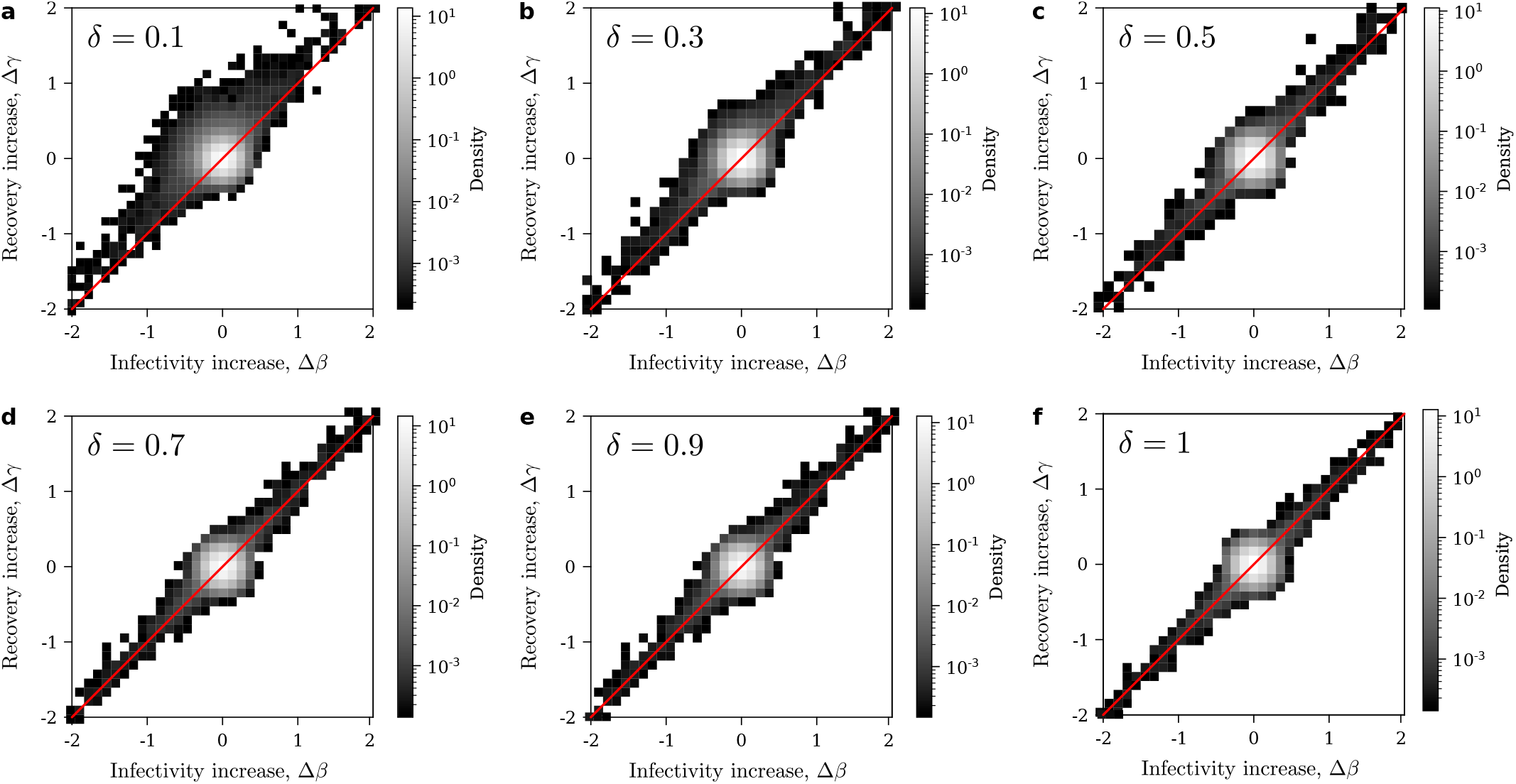
Joint probability density distribution of increases in model parameters *β* and *γ* between consecutive outbreaks. Death rates, **a** *δ* = 0.1, **b** 0.3, **c** 0.5, **d** 0.7, **e** 0.9, **f** 1. While the underlying random walk is perfectly symmetric (barring effects at the border of parameter space), cross-immune memory induces an asymmetry in the likelihood of observing increases in *β* and *γ* between consecutive successful strains. Symmetry is recovered as increased population renewal erases cross-immune memory from the population.

**SUP. FIG. 5.**
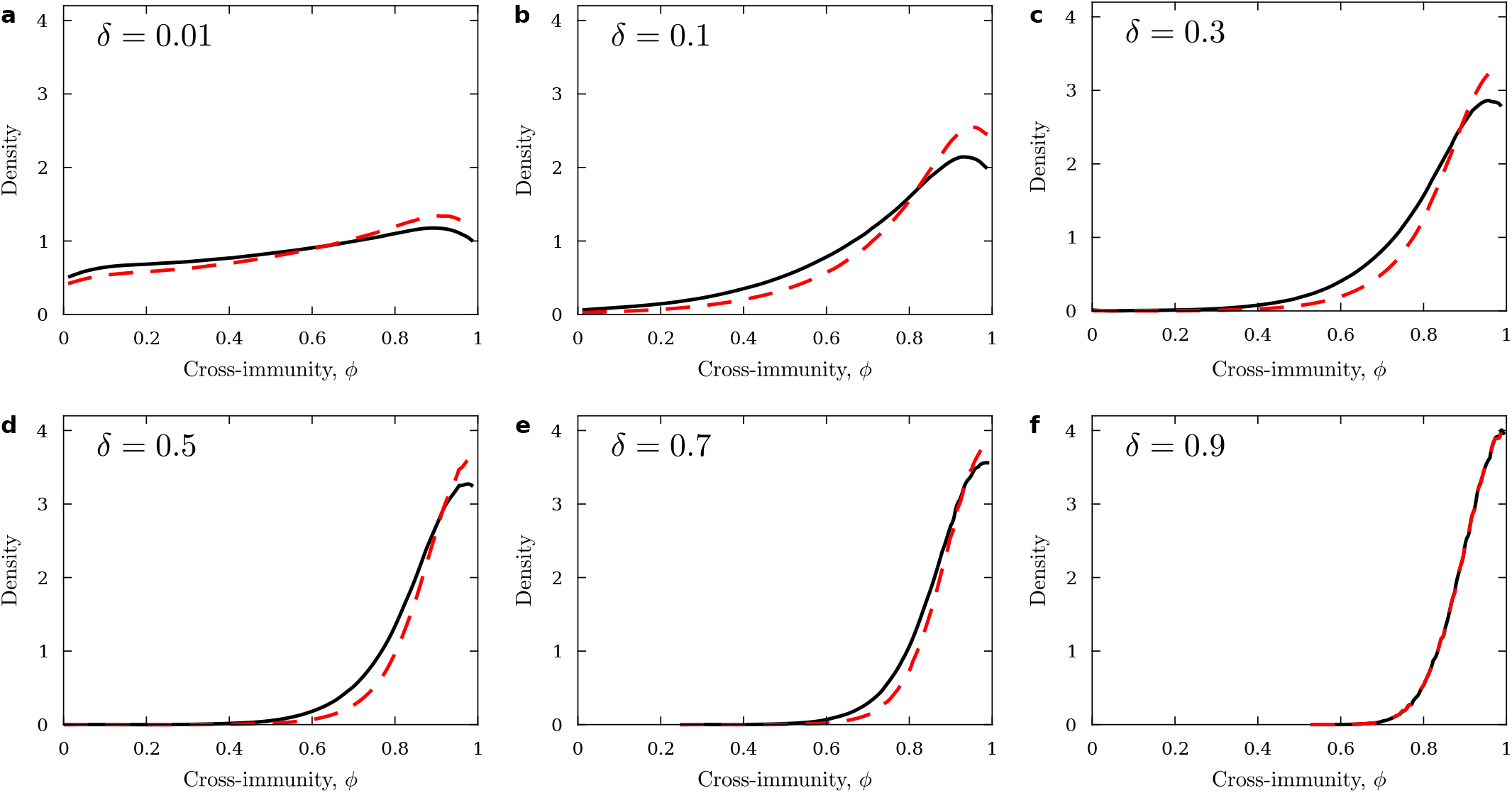
Probability density distributions of extant cross-immunity at outbreaks. Death rates, **a** *δ* = 0.1, **b** 0.3, **c** 0.5, **d** 0.7, **e** 0.9, **f** 1. In solid black, usual probability density distribution of extant cross-immunity at outbreaks. In dashed red, probability density distribution of extant cross-immunity at outbreaks weighting each *ϕ* by the amount of population that the ancient strain still offers protection to. For large memories, the distribution should approach the limit of an unbiased random walk (uniform except for border effects). For lower memory, cross-immunity cannot diffuse much from its starting point (*ϕ* = 1). The actual distribution has spikes at *ϕ* = 0 and *ϕ* = 1 due to the random walk persistently bouncing against the borders—this has been removed for clarity.

## Notes

### Competing Interest Statement

The authors have declared no competing interest.

